# What has been the impact of Covid-19 on Safety Culture? A case study from a large metropolitan teaching hospital

**DOI:** 10.1101/2020.06.15.20129080

**Authors:** Max Denning, Ee Teng Goh, Alasdair Scott, Guy Martin, Sheraz Markar, Kelsey Flott, Sam Mason, Jan Przybylowicz, Melanie Almonte, Jonathan Clarke, Jasmine Winter-Beatty, Swathikan Chidambaram, Seema Yalamanchili, Benjamin Yong-Qiang Tan, Abhiram Kanneganti, Viknesh Sounderajah, Mary Wells, Sanjay Purkayastha, James Kinross

## Abstract

**Introduction:** Covid-19 has placed an unprecedented demand on healthcare systems worldwide. A positive safety culture is associated with improved patient safety and in turn patient outcomes. To date, no study has evaluated the impact of Covid-19 on safety culture.

**Methods:** The Safety Attitudes Questionnaire (SAQ) was used to investigate safety culture at a large UK teaching hospital during Covid-19. Findings were compared with baseline data from 2017. Incident reporting from the year preceding the pandemic was also examined.

**Results:** Significant increased were seen in SAQ scores of doctors and ‘other clinical staff’, there was no change in the nursing group. During Covid-19, on univariate regression analysis, female gender, age 40-49 years, non-white ethnicity, and nursing job role were all associated with lower SAQ scores. Training and support for redeployment were associated with higher SAQ scores. On multivariate analysis, non-disclosed gender (−0.13), non-disclosed ethnicity (−0.11), nursing role (−0.15), and support (0.29) persisted to significance. A significant decrease (p<0.003) was seen in error reporting after the onset of the Covid-19 pandemic.

**Discussion:** This is the first study to report SAQ during Covid-19 and compare with baseline. Differences in SAQ scores were observed during Covid-19 between professional groups and compared to baseline. Reductions in incident reporting were also seen. These changes may reflect perception of risk, changes in volume or nature of work. High-quality support for redeployed staff may be associated with improved safety perception during future pandemics.

**WHAT IS ALREADY KNOWN ON THE SUBJECT:**

- Safety culture is associated with patient safety and outcomes
- This is the first study to investigate safety culture during the Covid-19 pandemic
- This study uses the Safety Attitudes Questionnaire (SAQ) and Datix incident reporting data to investigate determinants of safety climate during the Covid-19 pandemic.
- Safety climate is context specific, this study is strengthened by the availability of benchmarking data from before the onset of the Covid-19 pandemic.
- Significant differences in SAQ scores between professional groups were observed during Covid-19.
- Gender, ethnicity and job role were significant determinants of safety attitudes.
- Support during redeployment was associated with improved safety attitudes.
- The number of incidents that were reported reduced significantly during Covid-19, although the number of events leading to harm remained constant.

## INTRODUCTION

The provision of safe, high-quality care should always be an essential aim of a healthcare system, even during crises such as natural disasters, conflicts, or pandemics. To date, there has been no systematic evaluation of the impact of the Covid-19 pandemic on patient safety.

In addition to its effect on patients, Covid-19 has also placed a significant burden on healthcare systems, with drastic ramifications for the way healthcare is delivered. Rapid changes in models of care delivery were seen during the Covid-19 pandemic including increased workload, redeployment of staff to unfamiliar clinical environments, cancellation of routine services, and the requirement to treat patients suffering from a novel disease about which little was known. Working in these challenging conditions may impact on the ability of staff to deliver safe and effective care. Previous work has identified organizational breakdown, inadequate staffing, increased production pressures and provider fatigue as contributors to poor patient safety(1). Conceivably all these factors may have been present during the pandemic, and as such patient safety during the Covid-19 pandemic merits further investigation. A way of obtaining insight into the state of patient safety is via assessing safety culture and incident reporting.

Safety culture refers to “managerial and worker attitudes and values related to the management of risk and safety(2)” and is positively associated with both patient safety(3-5) and clinical outcomes(6, 7). Incident reporting is an important aspect of safety culture(8). Reporting systems do more than just measure harm at the organisational level; they identify system vulnerabilities, promote learning, and indicate willingness of staff to raise concerns.

### Aim

This study aims to assess institutional safety culture during the Covid-19 pandemic using a large London teaching hospital as a case study.

## METHODS

### Setting

Local institutional ethical approval was obtained (ICREC Ref:20IC5890). Imperial College Healthcare NHS Trust (ICHNT) is a group of 5 hospitals located in central London. The trust has over 1.2 million patient contacts per year. Over 12,000 staff work at the trust, of which 2,700 are doctors, 4,800 are nurses or midwives and 770 are allied health professionals. 52% are from Black, Asian or Mixed Ethnic (BAME) backgrounds.

In response to the escalating Covid-19 pandemic, changes included introduction of telemedicine outpatient clinics, mass usage of online off-site access for clinicians, cessation of elective surgical services, and expansion of adult critical care capacity by repurposing operating theatre recovery areas and paediatric intensive care beds. Additionally, significant changes were made to the workforce including redeployment to in-demand areas such as critical care or general medical wards. The nature and clinical characteristics of the Covid-19 caseload at ICHNT have been described elsewhere(9).

### Data collection

Baseline data collection was conducted using Survey Monkey (SurveyMonkey LLC, USA) between 12th October 2016 and 10th January 2017. Data for the Covid-19 group were completed using Google forms (Google LLC, USA) between 22nd April 2020 and 22nd May 2020. In both instances, invitations were sent by email to all staff, regardless of job role.

Two reminder emails were sent and 3 days of in-person convenience sampling was performed. Questionnaires were anonymous to encourage open responses.

### Questionnaire items

#### Demographic questions

The baseline questionnaire included questions related to job role, type of contract, and department (Appendix A). For the purpose of analysis, staff were categorised into doctors, nurses, and other clinical staff (including allied health professionals, healthcare support workers, pharmacists etc). During the Covid-19 data collection period, questions relating to personal characteristics were expanded to include: age, ethnicity, gender, and redeployment (Appendix B).

#### SAQ

The Safety Attitudes Questionnaire (SAQ) was used to assess perceptions of safety across six domains: safety climate, teamwork, stress recognition, perception of management, working conditions and job satisfaction. The SAQ is a reliable psychometric instrument(10) and is one of the most commonly used tools for measuring safety climate. It has been validated in a variety of settings including operating theatres, emergency care, intensive care, as well as medical and surgical wards(7). The SAQ is widely utilized in different countries and has been adapted for multiple different languages(7, 11-15). The tool is effective at comparing across locations, staff groups, and time. On all scales, higher scores represent a more positive perception of safety(10), which have been associated with positive clinical outcomes(16).

SAQ scores were calculated based on the 33 questions common to both surveys (Appendix C). Each question is associated with a given subscale or the overall score and includes a statement followed by a 5-point Likert scale from “*strongly disagree*” to “*strongly agree*”. Scores are expressed as the proportion of respondents that *“agree”* or “*strongly agree*” with a given statement. Vice versa for negatively worded questions. Questions left blank or answered “*not applicable*” are excluded from the analysis.

### Statistical Analysis

To achieve at least a precision of 5% on the SAQ a minimum sample size of 400 was required.

Group characteristics were demonstrated with simple descriptive statistics using Pearson chi-square tests to compare the distribution of characteristics among groups. Data were analysed for normal distribution using the Shapiro-Wilk test. SAQ data were found to be non-parametric, so Kruskal-Wallis tests were used to compare SAQ across groups. Violin plots comparing SAQ scores among professions were created using RStudio v1.2 (Version 1.2.5042. 2020 RStudio, Inc).

For the Covid-19 cohort, univariate quantile regression was performed to identify which characteristics influence SAQ score and the direction of any effect. Factors statistically associated with SAQ score in the univariate analysis (ethnicity, role, training, support) were assessed further in the multivariate model using a backwards stepwise approach. The final multivariate model controlled for factors that were significant or deemed important to control for (age, gender, ethnicity, role, redeployment). Redeployed staff were analysed as a subgroup to identify the impact of training and support in relation to redeployment. Confidence intervals were obtained by using 20 bootstrap samples. Statistical significance was set at p ≤ 0.05. Data were analysed using Stata v14 (StataCorp. 2015. Stata Statistical Software: Release 14. College Station. TX: StataCorp LP).

### Incident reporting

Incident reporting is another important indicator of safety climate and willingness to learn safety lessons. There is extensive research demonstrating the benefits of reporting.

Incidents reported through the Datix reporting system (Datix Limited, UK) between 01/04/19 and 10/05/20 were examined. These were classified as resulting in “harm”, “no harm”, or “near misses(17)”. These were plotted using a statistical process control (SPC) XmR chart. SPC charts are a tool used to graphically depict and statistically analyse incident reporting, allowing users to identify whether changes are due to natural (common cause) variation, or special cause variation. A detailed explanation of the methodology for plotting SPC charts can be found elsewhere(18). The upper and lower confidence intervals were set at 3 standard deviations based on the first 20 observations in order to define inherent variation in the process(18). The process mean and control limits were then recalculated when 8 consecutive points fell above or below the control line(18).

## RESULTS

At baseline, there were 1,580 responses. This included 166 doctors, 616 nurses, 374 other clinical staff. Nonclinical staff (424) and those that filled in none of the 33 SAQ questions (117) were excluded. The median overall SAQ score at baseline was 0.70.

During Covid-19, 455 responses were received. This included 183 nurses,105 doctors and 127 other clinical staff. 40 non-clinical staff responded, who were excluded from the analysis. Respondent characteristics for both groups are shown in Table 1. The median overall SAQ score was 0.73.

**Table 1.**
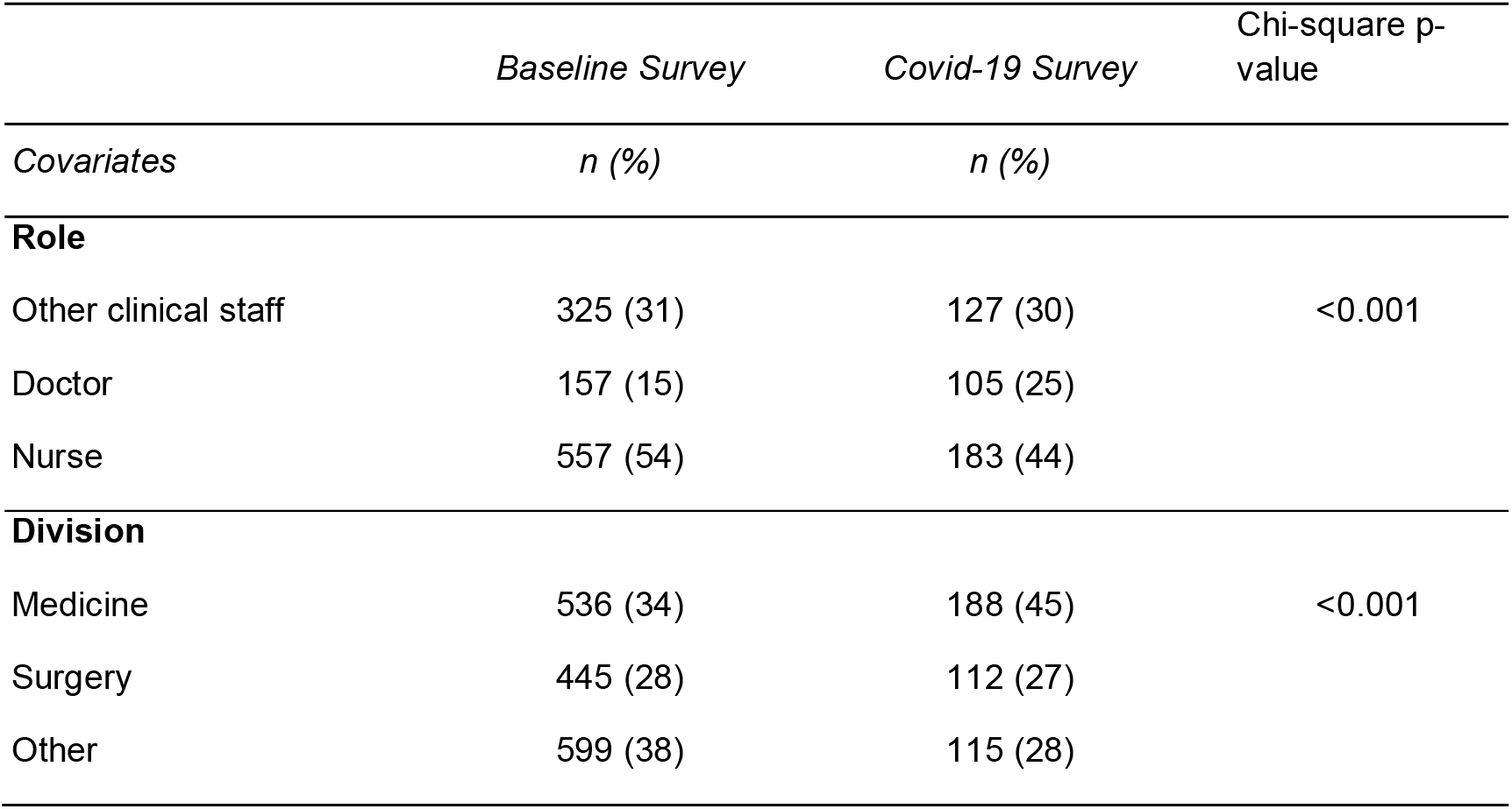
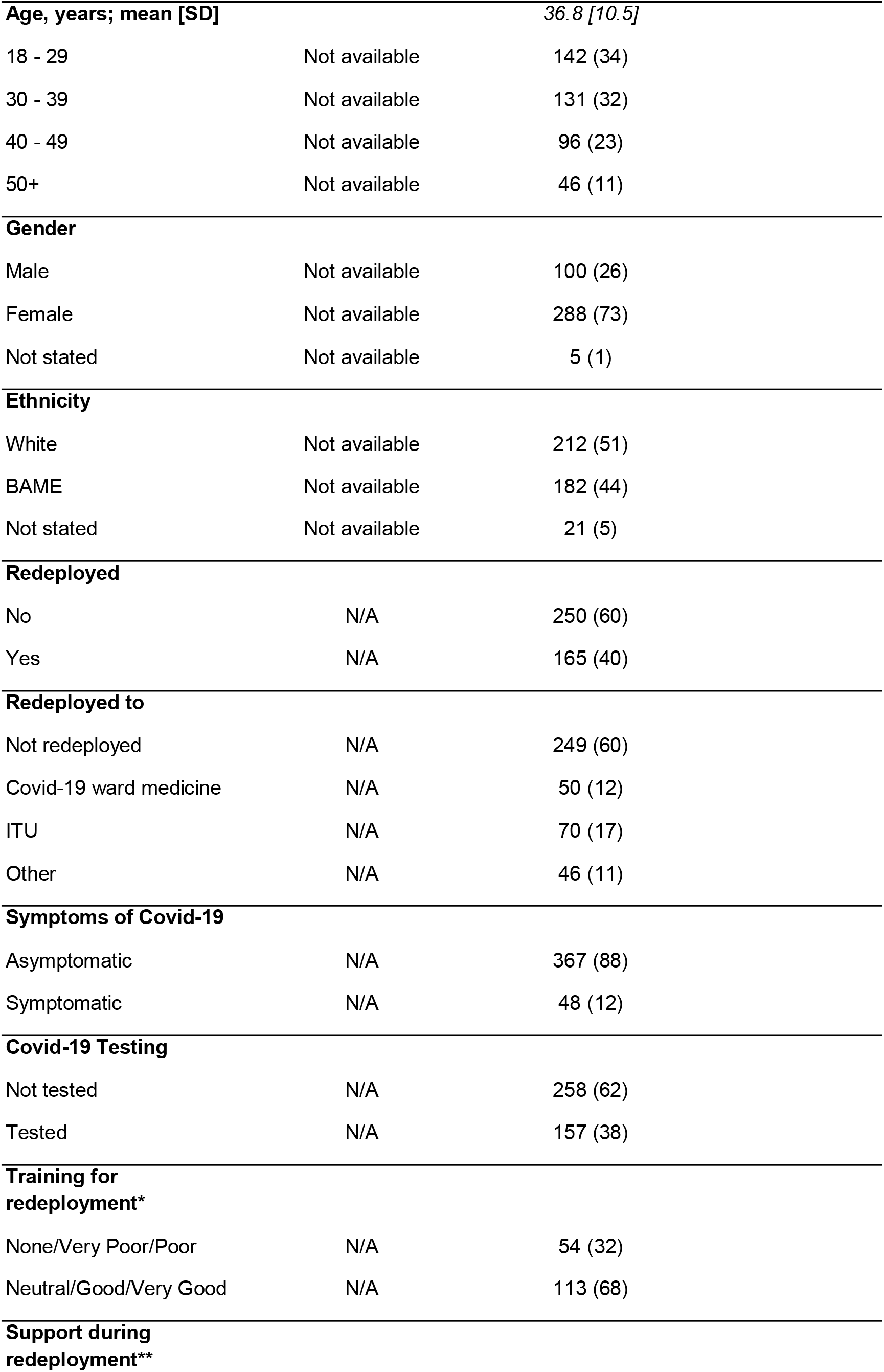
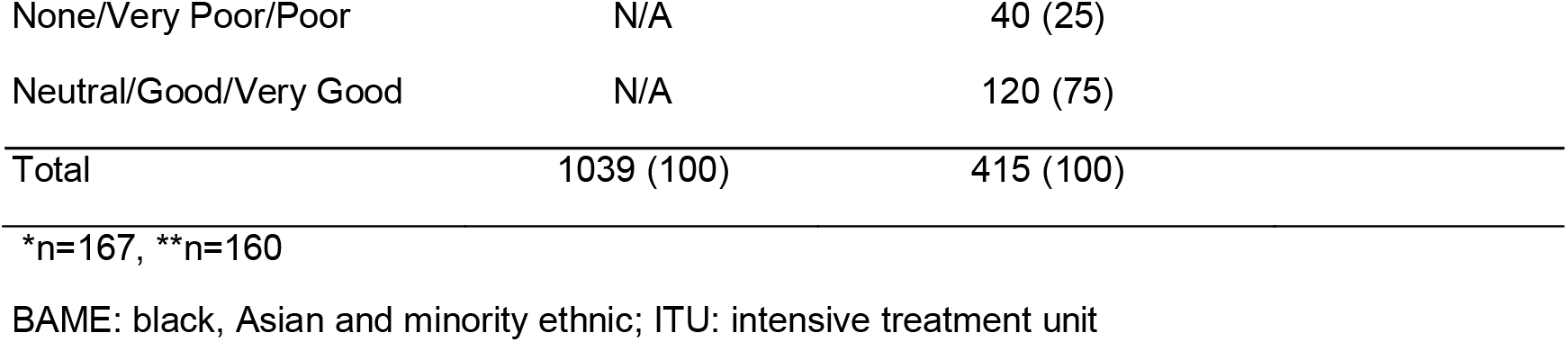
Respondent Characteristics

|  | *Baseline Survey* | *Covid-19 Survey* | Chi-square p-value |
| --- | --- | --- | --- |
| *Covariates* | *n (%)* | *n (%)* |  |
| **Role** |  |  |  |
| Other clinical staff | 325 (31) | 127 (30) | <0.001 |
| Doctor | 157 (15) | 105 (25) |  |
| Nurse | 557 (54) | 183 (44) |  |
| **Division** |  |  |  |
| Medicine | 536 (34) | 188 (45) | <0.001 |
| Surgery | 445 (28) | 112 (27) |  |
| Other | 599 (38) | 115 (28) |  |

|  |  |  |
| --- | --- | --- |
| **Age, years; mean [SD]** |  | 36.8 [10.5] |
| 18 - 29 | Not available | 142 (34) |
| 30 - 39 | Not available | 131 (32) |
| 40 - 49 | Not available | 96 (23) |
| 50+ | Not available | 46 (11) |
| **Gender** |  |  |
| Male | Not available | 100 (26) |
| Female | Not available | 288 (73) |
| Not stated | Not available | 5 (1) |
| **Ethnicity** |  |  |
| White | Not available | 212 (51) |
| BAME | Not available | 182 (44) |
| Not stated | Not available | 21 (5) |
| **Redeployed** |  |  |
| No | N/A | 250 (60) |
| Yes | N/A | 165 (40) |
| **Redeployed to** |  |  |
| Not redeployed | N/A | 249 (60) |
| Covid-19 ward medicine | N/A | 50 (12) |
| ITU | N/A | 70 (17) |
| Other | N/A | 46 (11) |
| **Symptoms of Covid-19** |  |  |
| Asymptomatic | N/A | 367 (88) |
| Symptomatic | N/A | 48 (12) |
| **Covid-19 Testing** |  |  |
| Not tested | N/A | 258 (62) |
| Tested | N/A | 157 (38) |
| **Training for redeployment\*** |  |  |
| None/Very Poor/Poor | N/A | 54 (32) |
| Neutral/Good/Very Good | N/A | 113 (68) |
| **Support during redeployment\*\*** |  |  |
| None/Very Poor/Poor | N/A | 40 (25) |
| Neutral/Good/Very Good | N/A | 120 (75) |
| Total | 1039 (100) | 415 (100) |

\*n=167, \*\*n=160
BAME: black, Asian and minority ethnic; ITU: intensive treatment unit

### Comparison

Distribution of SAQ scores by professional group is demonstrated in Figure 1. The overall SAQ score improved for all respondents from 0.70 to 0.73 (p<0.05). Comparison of SAQ subscales at baseline and during Covid-19 are shown in Table 2. Significant changes were seen in other clinical staff (p<0.001) and doctors (p<0.001). There was no significant change in score for the nursing group (p>0.05).

**Table 2.**
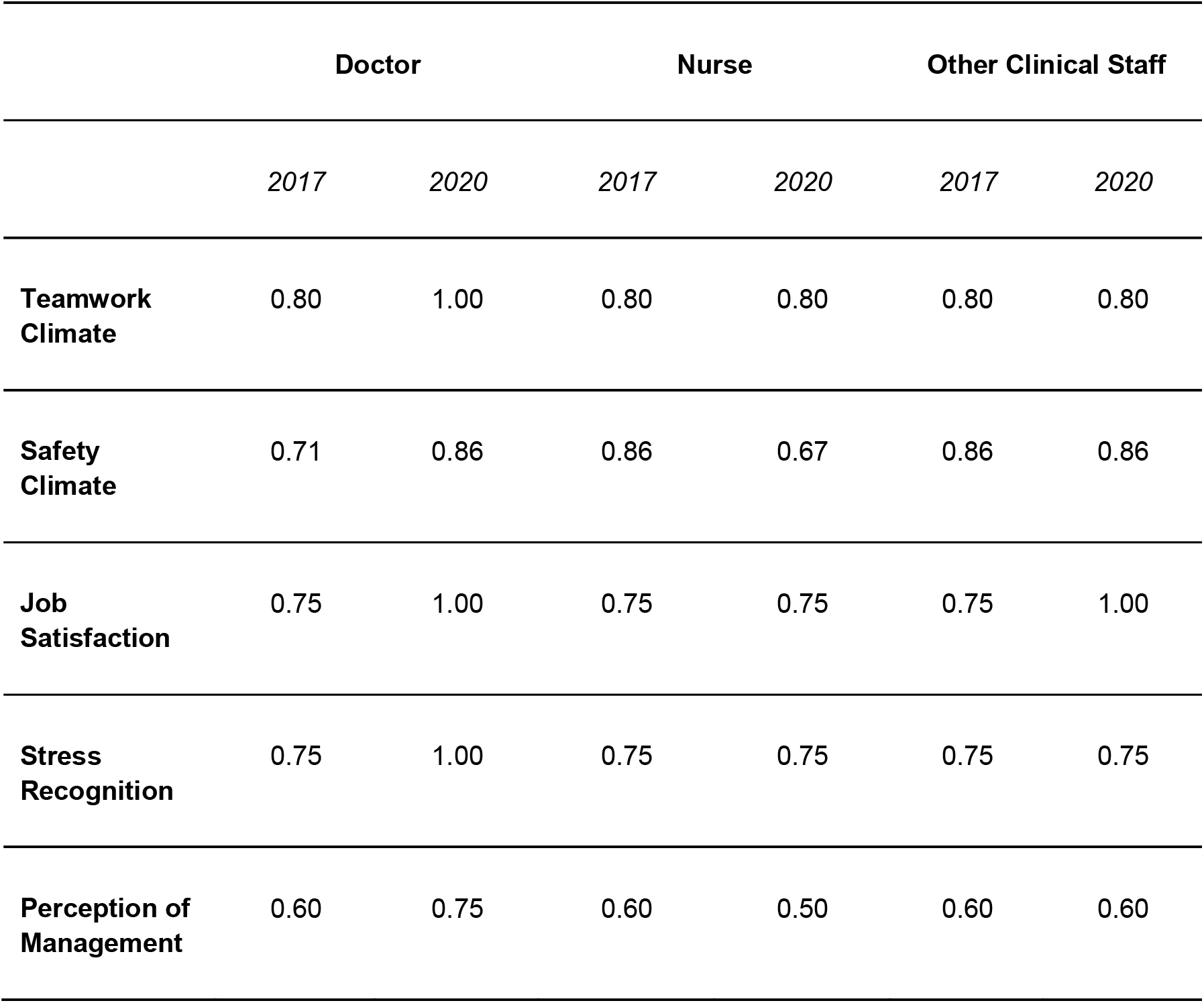
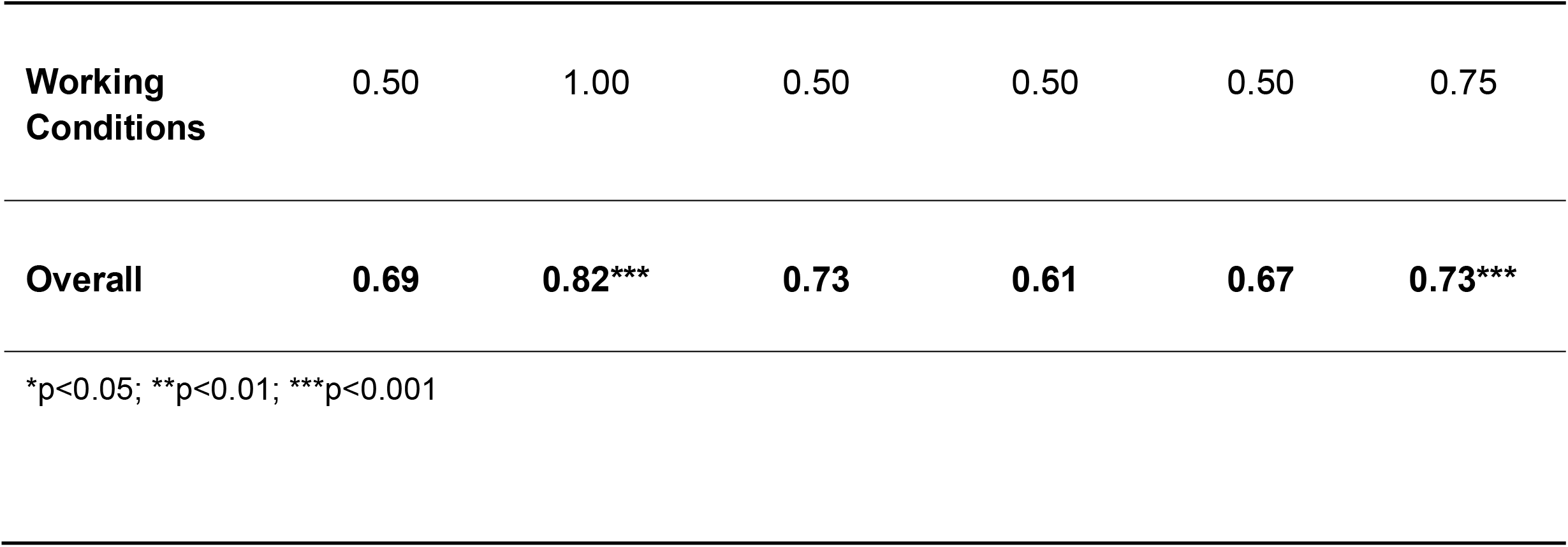
Median SAQ Scores before and during Covid-19

|  | Doctor |  | Nurse |  | Other Clinical Staff |  |
| --- | --- | --- | --- | --- | --- | --- |
|  | 2017 | 2020 | 2017 | 2020 | 2017 | 2020 |
| **Teamwork Climate** | 0.80 | 1.00 | 0.80 | 0.80 | 0.80 | 0.80 |
| **Safety Climate** | 0.71 | 0.86 | 0.86 | 0.67 | 0.86 | 0.86 |
| **Job Satisfaction** | 0.75 | 1.00 | 0.75 | 0.75 | 0.75 | 1.00 |
| **Stress Recognition** | 0.75 | 1.00 | 0.75 | 0.75 | 0.75 | 0.75 |
| **Perception of Management** | 0.60 | 0.75 | 0.60 | 0.50 | 0.60 | 0.60 |
| **Working Conditions** | 0.50 | 1.00 | 0.50 | 0.50 | 0.50 | 0.75 |
| **Overall** | **0.69** | **0.82\*\*\*** | **0.73** | **0.61** | **0.67** | **0.73\*\*\*** |

\*p<0.05; \*\*p<0.01; \*\*\*p<0.001

**Figure 1.**
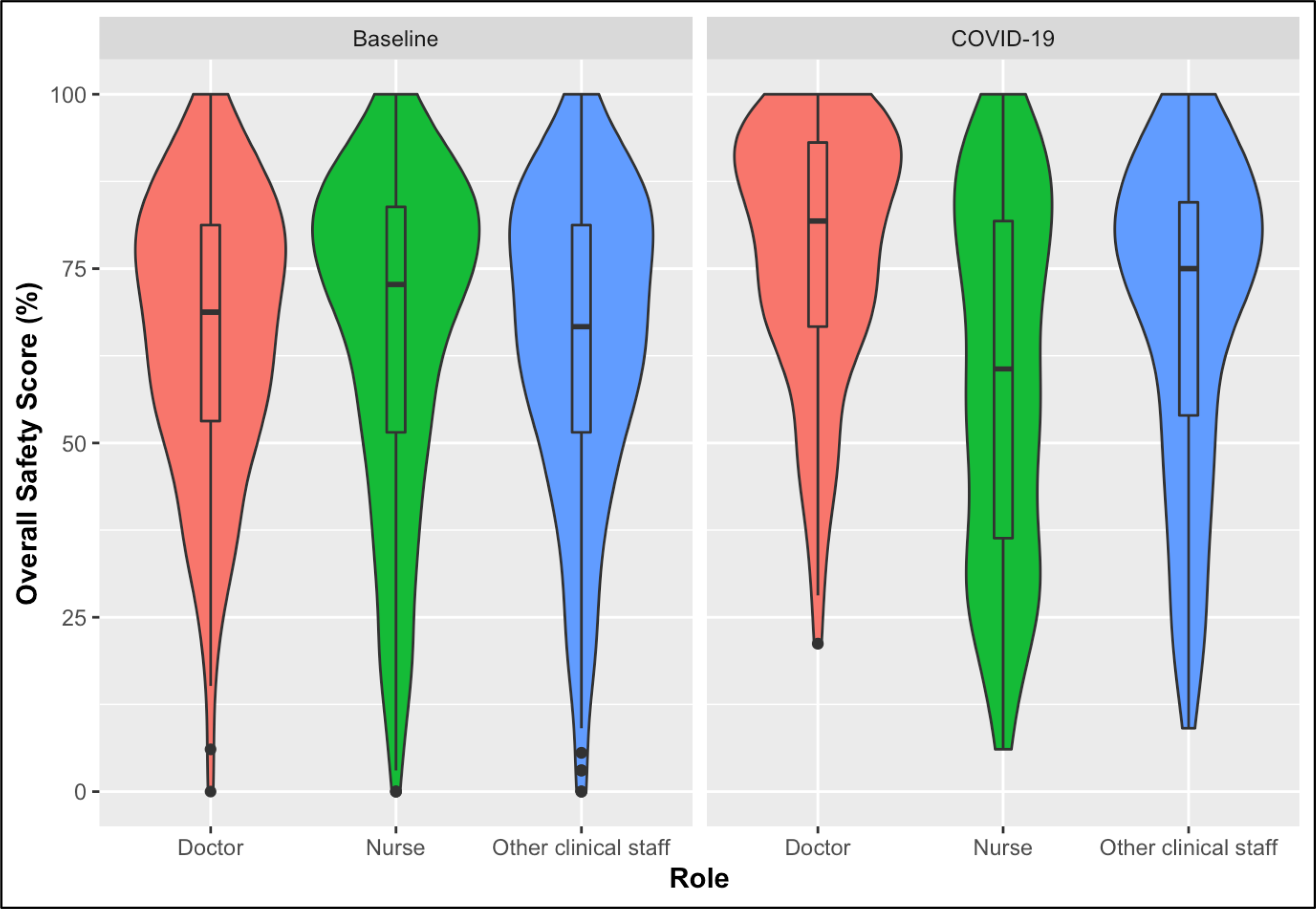
Violin plot demonstrating SAQ scores at baseline and during Covid-19, by profession. Width represents the proportion of respondents with a given score. Central line represents the median. Box represents interquartile range. Lines represent the range of scores. Dots represent outliers (more than 1.5 times the interquartile range)

### Regression Analysis

On univariate analysis of the responses from the Covid-19 cohort, age 40-49 years (p<0.01), non-disclosed gender (p<0.001), BAME ethnicity (p<0.001), nursing job role (p<0.001), other clinical staff job role (p<0.01) were predictors of lower SAQ scores. On subgroup analysis of those redeployed, age 40-49 (p<0.05), BAME ethnicity (p<0.01), nursing job role (p<0.01) and having symptoms (p<0.01) predicted lower SAQ scores. Training for redeployment (p<0.001) and support during redeployment (p<0.001) predicted higher SAQ scores.

On multivariate analysis, controlling for all other covariates in the model, non-disclosed gender (−0.13, −0.26 − 0.00), non-disclosed ethnicity (−0.11, −0.22 − 0.00) and nursing role (−0.15, −0.24 − 0.06), remained significant predictors of low SAQ scores. In the redeployed subgroup (n=139), support (0.29, 0.14 − 0.44) and nursing role (−0.12, −0.23 −-0.01) persisted to significance. Detailed results of the quantile regression analysis are depicted in Table 3.

**Table 3.**
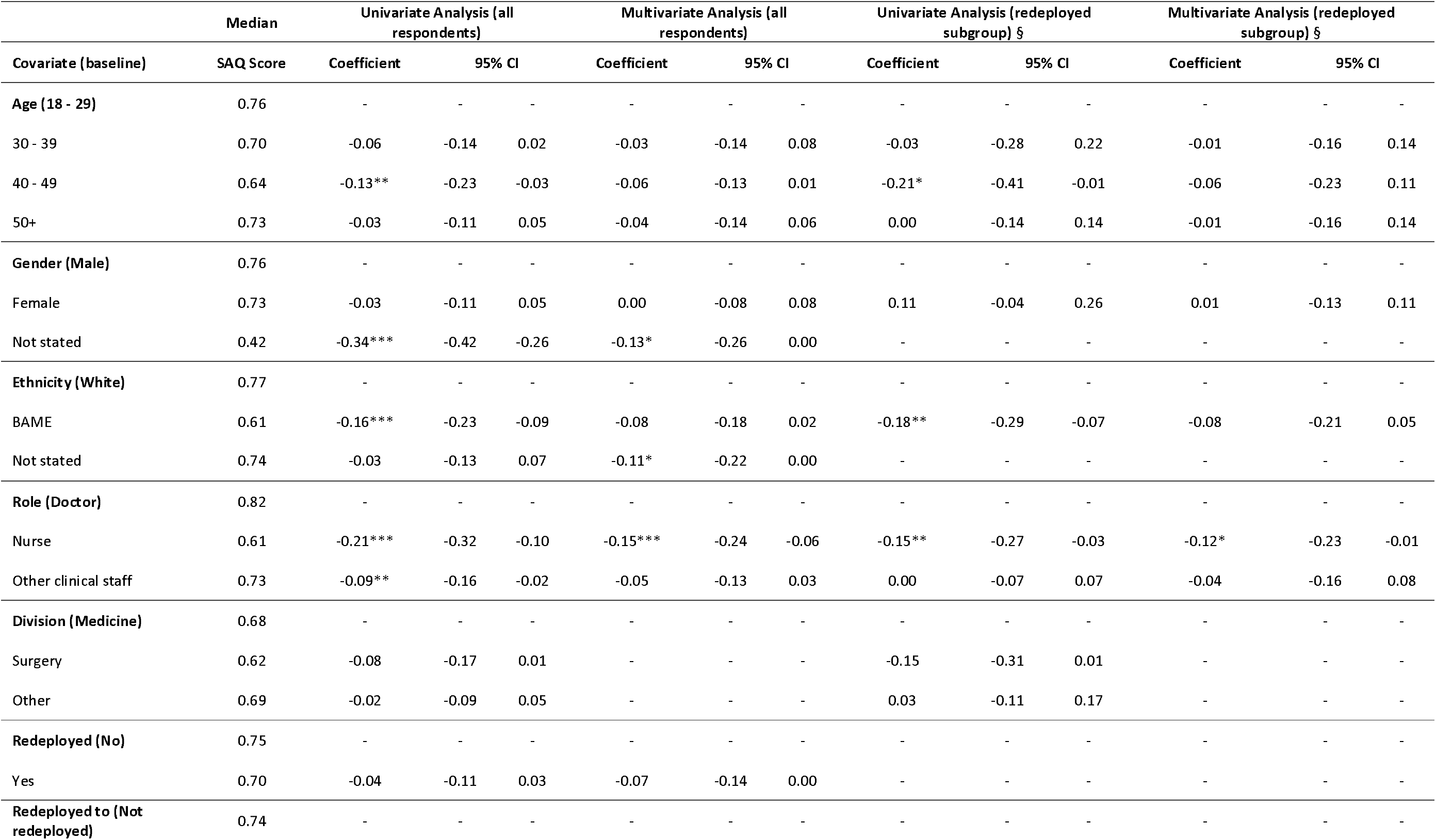
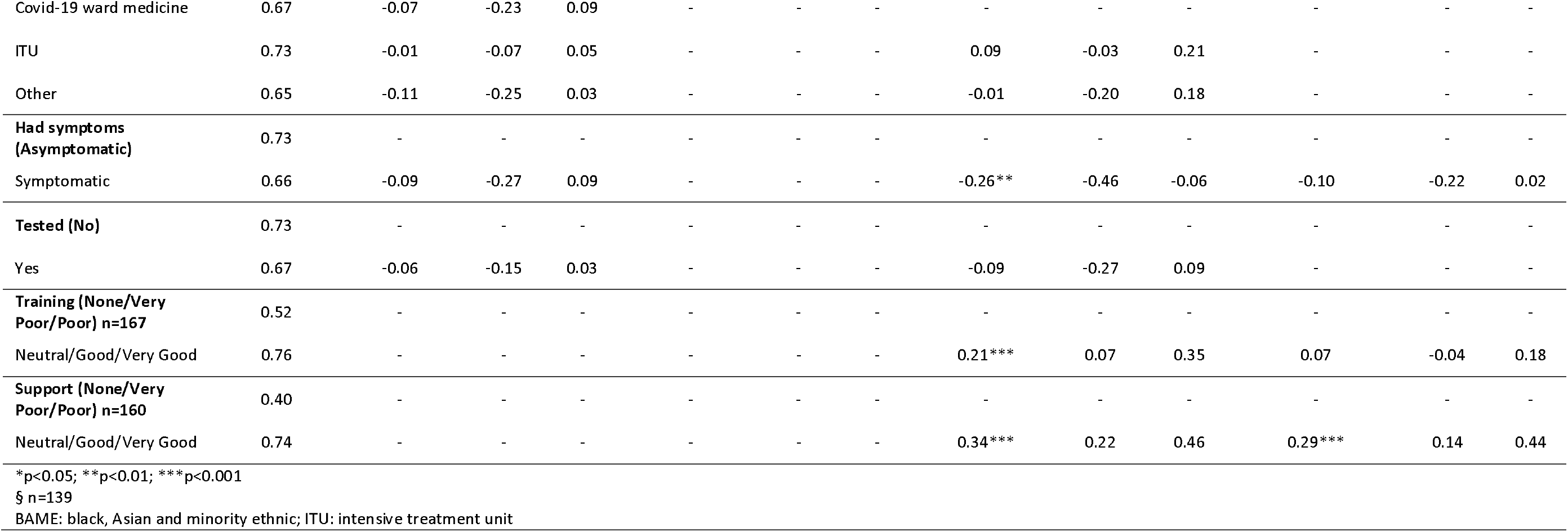
Quantile regression of SAQ scores during the Covid-19 pandemic

### Error reporting

Weekly patient safety incidents are demonstrated as an XmR Control chart in figure 2. Following the arrival of the first Covid-19 case in the trust, error reporting significantly reduced. In the 8 weeks following the first Covid-19 patient arriving at the trust, the number of weekly error reports consistently fell below the 52-week mean. On 6 of the 8 weeks, the rate was more than 3 standard deviations below the weekly mean. When broken down by type, ‘No Harm’ and ‘Near Miss’ reports significantly reduced (p<0.003). However, the number of ‘Harm’ incidents reported did not significantly change.

**Figure 2.**
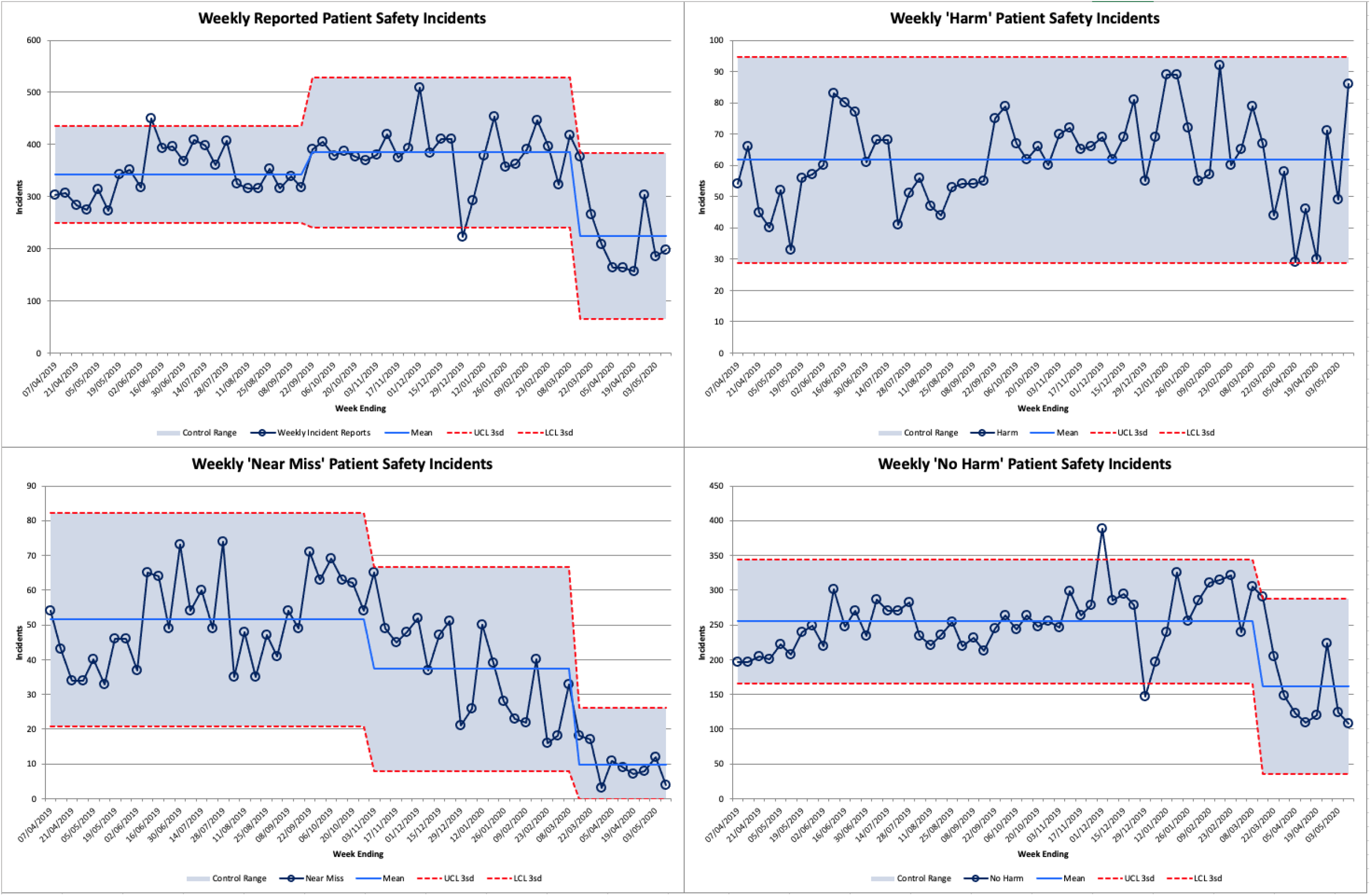
Incident Reporting Year to Date

## DISCUSSION

To the authors’ knowledge, this is the first study investigating safety culture during the Covid-19 pandemic. This analysis has shown significantly higher SAQ scores in doctors and other clinical staff compared to baseline, but no change for nurses. Due to changes in the clinical context, a direct comparison of the two samples may be subject to limitations. However, comparisons between groups at each time point will still provide valuable insights. During Covid-19, nurses demonstrated significantly lower SAQ scores compared with doctors and other clinical staff. This contrasts with the baseline cohort where nurses had the highest safety attitude scores. In the Covid-19 cohort, whilst controlling for other covariates, non-disclosure of gender, non-disclosure of ethnicity and the nursing job role were predictive of poor SAQ score. In contrast, provision of support was predictive of high SAQ score.

The results may reflect the unequal accessibility and effectiveness of current efforts to support stuff. There have been both national and local initiatives to support staff during the Covid-19 pandemic such as access to free food, staff counselling services, and transport allowances. Nationally, the ‘clap for carers’ campaign, commercial discounts, and social media initiatives may have also contributed. Other possible explanations include a sense of solidarity in which multiple staff came together to overcome the challenge of a novel disease. The lower scores for working conditions and job satisfaction in the nursing staff group compared to other roles may suggest that not all staff had equal access to support initiatives or that such initiatives were not equally effective for all staff.

The finding that staff who did not disclose gender or ethnicity had lower SAQ scores, may reflect a wish to avoid being identified for negative views or responses on a survey, especially on questions relating to perceptions of management. That being said, this group of respondents were very few in number and as such the true significance of this finding is unclear.

Environmental factors, working conditions, cognitive load, stress, and fatigue are all factors that impact on individual performance and may have differentially affected staff during the Covid-19 pandemic. Safety climate and perception of management decreased in nurses, which may reflect experiences of topical issues such as provision of personal protective equipment provision or testing. Staff groups spend different amounts of time with Covid-19 patients and therefore may have differing requirements, concerns and experiences of such resources or initiatives. Interestingly, there was no significant association between staff testing and SAQ score.

Support during redeployment was associated with significantly higher SAQ scores. This suggests that in addition to the intended effects of improving staff wellbeing, support programmes may be associated with improved perceptions of safety. However, a prospective design would be needed to demonstrate causality. The specific aspects of support perceived positively by staff also warrant further investigation.

### Incident reporting

There was a significant reduction in the overall rate of incident reporting following the onset of Covid-19. However, no statistically significant change was seen in the number of incidents reporting patient ‘harm’. Incident reporting is well recognised as an indicator of safety culture(19), however it is unclear if that association would persist under conditions of the pandemic.

This reduction in reporting may be explained by: staff having less time to file incident reports due to increased workload, a change in the perceived relative importance of error reporting, or fewer patient safety incidents occurring due to changes in the nature of work, such as the cessation of routine services. Irrespective of aetiology, the reduction in incident reporting represents potential lost opportunities to learn and improve safety, which is of importance at such an unprecedented time for service delivery.

### Recommendations for future pandemics

Further information is needed to understand why personal characteristics were associated with perceptions of safety. However, through well designed and targeted support, it may be possible to mitigate some of these differences.

Organisational factors such as shift patterns, redeployment opportunities, and occupational risk management may represent targets for interventions. Besides that, incident reporting is an area of particular importance during a change in service provision, as this provides opportunities to understand what is impacting on safety in a particular healthcare context. Initiatives should therefore be considered that facilitate staff to incident report even during pandemics.

Quality improvement and patient safety teams can support the pandemic response. Gurses et al(20) have described the importance and potential benefits of using human factors experts as part of a pandemic response. The group suggested that integration of human factors principles could help “design, adapt and reconfigure work systems, maximize individual and team performance under high-risk, high-stakes environments, while minimizing the introduction of new significant safety risks or unintended consequences into the work system”. Staines et al have proposed a five-step strategy to support organisations responses during the Covid-19 pandemic(21). Steps proposed place emphasis on clear communication, just-in-time guidelines and training, adequate support for patient and staff wellbeing, improving operational flow, and risk management. The strategy provides a framework for rapid implementation should further peaks of the Covid-19 pandemic occur and can be utilized in other pandemics or protracted major incidents. Such initiatives may contribute to improved safety culture.

### Limitations

Both surveys had a limited response amongst staff introducing a possible response bias. This was more pronounced during the Covid-19 sample (5% compared with 15%). Although the response rate was low, the absolute number of respondents represented a large number and diverse range of staff. Prior research has highlighted the the poor response rates common in surveys of clinicians(22). This may have been accentuated by changes in the nature of work, increased workload, time pressure, or survey fatigue during the Covid-19 pandemic. In instances where response rates are likely to be low, previous work(23) has highlighted the importance of collecting data using a combination of methods. To address potential response bias, we made use of both email invitations and in-person sampling to capture responses from individuals that might not have otherwise participated.

The clinical context and workforce were different at the two points in time. Whilst differences between professional groups at either point in time are likely to be valid, drawing inferences regarding the changes in absolute scores over time is subject to limitations. Nevertheless, the sample was from a large teaching hospital with a diverse workforce, and our sample was representative of the underlying workforce demographics. In the Covid-19 cohort, for example, 73% of respondents were female. Broadly in line with the NHS workforce, which is 77% female(244).

The categories and rate of incident reporting have been described; however, the underlying incidents have not been analysed. It is therefore not possible to identify whether the decrease in incident reporting suggests changing safety culture, attitudes about reporting or a change in the underlying nature of work.

Finally, this study investigates safety attitudes which, while providing valuable insight, may not provide the complete picture on patient safety during the Covid-19 pandemic. Further research on clinical outcomes during this period is warranted.

## CONCLUSION

This is the first study to investigate safety culture during the Covid-19 pandemic. Using a cross-sectional survey-based design alongside incident data, we have identified significant differences in safety attitudes before and during the Covid-19 pandemic. Within the Covid-19 cohort, nursing role was associated with lower SAQ scores as compared to other roles. Support was associated with better SAQ scores. The rate of incident reporting reduced following the onset of the pandemic. These findings underline the importance of targeted high-quality support during future pandemics.

## Data Availability

The data generated during and/or analysed during the current study are available from the corresponding author upon reasonable request.

## Notes

**Competing interests** No conflicts to declare

**Funding** No funding was received in support of this paper. Johnson and Johnson have supported the activities of the PanSurg collaborative with an educational grant.

### Competing Interest Statement

The authors have declared no competing interest.

### Clinical Trial

Observational study, not a prospective clinical trial

### Clinical Protocols

https://drive.google.com/file/d/1Odpo1iASbNKS8od4NrmEkh6q2QjpFHfX/view

### Funding Statement

No funding was received in support of this paper. Johnson and Johnson have supported the activities of the Pansurg collaborative with an educational grant.

### Author Declarations

Imperial College Research Ethics Committee Reference:20IC5890

## References

1. Jha AK, Prasopa-Plaizier N, Larizgoitia I, et al. Patient safety research: an overview of the global evidence. Quality and Safety in Health Care 2010;19(1):42–7.

2. Flin R, Winter J, Sarac C, et al. Human Factors in Patient Safety: Review of Topics and Tools. Geneva: 2009.

3. Hofmann D, Mark B. An Investigation Of The Relationship Between Safety Climate And Medication Errors As Well As Other Nurse And Patient Outcomes. Personnel Psychology 2006;59(4):847–69.

4. Naveh E, Katz-Navon T, Stern Z. Treatment Errors in Healthcare: A Safety Climate Approach. Management Science 2005;51(6):948–60.

5. Zohar D, Livne Y, Tenne-Gazit O, et al. Healthcare climate: a framework for measuring and improving patient safety. Critical care medicine 2007;35(5):1312–7.

6. Murphy T. CCHSA Client/Patient Safety Culture Assessment Project: lessons learned. Healthcare quarterly (Toronto, Ont) 2006;9(2):52-4, 2.

7. The Evidence Centre. Evidence Scan: Measuring Safety Culture. The Health Foundation, 2011.

8. Institute of Medicine Committee on Quality of Health Care in A In: Kohn LT, Corrigan JM, Donaldson MS, editors. To Err is Human: Building a Safer Health System. Washington (DC): National Academies Press; 2000.

9. Perez Guzman P, Daunt A, Mukherjee S, et al. Clinical Characteristics and Predictors of Outcomes of Hospitalized Patients with COVID-19 in a London NHS Trust: A Retrospective Cohort Study. London: Imperial College Healthcare NHS Trust, 2020 29 April. Report No.: 17.

10. Sexton JB, Helmreich RL, Neilands TB, et al. The Safety Attitudes Questionnaire: psychometric properties, benchmarking data, and emerging research. BMC health services research 2006;6:44.

11. Deilkås ET, Hofoss D. Psychometric properties of the Norwegian version of the Safety Attitudes Questionnaire (SAQ), Generic version (Short Form 2006). BMC health services research 2008;8:191.

12. Kaya S, Barsbay S, Karabulut E. The Turkish version of the safety attitudes questionnaire: psychometric properties and baseline data. Quality & safety in health care 2010;19(6):572–7.

13. Lee WC, Wung HY, Liao HH, et al. Hospital safety culture in Taiwan: a nationwide survey using Chinese version Safety Attitude Questionnaire. BMC health services research 2010;10:234.

14. Nguyen G, Gambashidze N, Ilyas SA, et al. Validation of the safety attitudes questionnaire (short form 2006) in Italian in hospitals in the northeast of Italy. BMC health services research 2015;15:284.

15. Nordén-Hägg A, Sexton JB, Kälvemark-Sporrong S, et al. Assessing safety culture in pharmacies: the psychometric validation of the Safety Attitudes Questionnaire (SAQ) in a national sample of community pharmacies in Sweden. BMC clinical pharmacology 2010;10:8.

16. Pronovost P, Sexton B. Assessing safety culture: guidelines and recommendations. Quality and Safety in Health Care 2005;14(4):231–3.

17. Vincent C, Burnett S, Carthey J. The Measurement and Monitoring of Safety. London: Centre for Patient Safety and Service Quality Imperial College London, 2013 April. Report No.

18. Benneyan JC, Lloyd RC, Plsek PE. Statistical process control as a tool for research and healthcare improvement. Quality & safety in health care 2003;12(6):458–64.

19. Kagan I, Barnoy S. Organizational safety culture and medical error reporting by Israeli nurses. Journal of nursing scholarship: an official publication of Sigma Theta Tau International Honor Society of Nursing 2013;45(3):273–80.

20. Gurses AP, Tschudy MM, McGrath-Morrow S, et al. Overcoming COVID-19: What can human factors and ergonomics offer? Journal of Patient Safety and Risk Management 2020;25(2):49–54.

21. Staines A, Amalberti R, Berwick DM, et al. COVID-19: patient safety and quality improvement skills to deploy during the surge. International Journal for Quality in Health Care 2020.

22. Cook JV, Dickinson HO, Eccles MP. Response rates in postal surveys of healthcare professionals between 1996 and 2005: an observational study. BMC health services research 2009;9:160.

23. McMahon SR, Iwamoto M, Massoudi MS, et al. Comparison of e-mail, fax, and postal surveys of pediatricians. Pediatrics 2003;111(4 Pt 1):e299–303.

24. NHS Digital. Narrowing of NHS gender divide but men still the majority in senior roles Leeds: NHS Digital; 2018 [updated 29 June 2018; cited 2020 11 June]. Available from: https://digital.nhs.uk/news-and-events/latest-news/narrowing-of-nhs-gender-divide-but-men-still-the-majority-in-senior-roles.

